# Staphylococcal superantigens promote bacterial persistence following postoperative surgical site infection

**DOI:** 10.1101/2024.07.23.24310826

**Authors:** Karine Dufresne, Stephen W. Tuffs, Nicholas R. Walton, Katherine J. Kasper, Ivor Mohorovic, Farah Hasan, Tracey Bentall, David E. Heinrichs, Johan Delport, Tina S. Mele, John K. McCormick

**Affiliations:** Department of Microbiology and Immunology, University of Western Ontario, London, Canada; Department of Biochemistry and Microbiology, University of Victoria, Victoria, Canada; Division of Critical Care Medicine, Department of Medicine, University of Western Ontario, London, Canada; Department of Pathology, University of Western Ontario, London, Canada; Division of General Surgery, Department of Surgery, London Health Sciences Centre, University Hospital, London, Canada

**Keywords:** *Staphylococcus aureus*, superantigen, surgical site infection, bacteremia

## Abstract

*Staphylococcus aureus* is a predominant cause of postoperative surgical site infections and persistent bacteremia. Here we describe a patient that following a total knee arthroplasty subsequently experienced three episodes of *S. aureus* bacteremia over a period of 4 months. The initial blood stream isolate (SAB-0429) was a clonal complex (CC) 5 and methicillin resistant *S. aureus* (MRSA), whereas two subsequent blood stream isolates (SAB-0485 and SAB-0495) were CC5 isolates but methicillin sensitive *S. aureus* (MSSA). The two latter isolates harbored a plasmid encoding three superantigen genes not present in the primary MRSA isolate. SAB-0485 and SAB-0495 expressed the plasmid encoded staphylococcal enterotoxin R (SER) exotoxin and demonstrated increased superantigen activity compared with SAB-0429. Compared to SAB-0429, the latter isolates also demonstrated an increased bacterial burden in a mouse bacteremia model that was dependent on increased IFNγ production. Curing of the plasmid from SAB-0485 reduced this virulence phenotype. These findings suggest that the superantigen exotoxins may provide a selective advantage in chronic postsurgical infections.

## Introduction

*Staphylococcus aureus* is a common human colonizer and opportunistic pathogen capable of causing a wide array of infections that can range from superficial skin lesions to invasive infections including endocarditis, osteomyelitis and bacteremia. Fatality rates for *S. aureus* bacteremia (SAB) in both community and hospital settings are approximately 20-30% [1,2] and methicillin-resistant *S. aureus* (MRSA) strains are of major concern as treatment with antibiotics often fail to clear the bacteria within the patients until a correct regimen is found [3]. *S. aureus* is also an important cause of post-surgical site infections (SSIs) where infection due to MRSA has been related to a 7-fold increased risk of death, a 35-fold increased risk of hospital re-admission, and over three weeks of additional hospitalization [4]. When compared to matched control cases, orthopedic SSIs prolong hospital stays for an average of 2 weeks, approximately doubling the rates of re-hospitalization, increasing hospital costs by ∼300% [5,6]. Orthopedic SSI can also result in a decreased quality of life due to increased physical limitations from the infection [5]. For these reasons, *S. aureus* decolonization strategies of the orthopedic surgical team and of patients have been implemented and demonstrate a reduction of the total number of surgical site infections [7–9].

Both MRSA and methicillin-sensitive *S. aureus* (MSSA) can survive and persist during infection by manipulating the host using multiple virulence factors [10], including a unique family of toxins called superantigens (SAgs) [11]. SAgs trigger large scale activation of T cells independent of antigen presentation and this aberrant activation can lead to an overwhelming cytokine storm disease known as toxic shock syndrome [12]. During experimental bloodstream infection, *S. aureus* can survive and proliferate in organs including the liver and kidney and, in this context, staphylococcal SAgs can dramatically enhance bacterial burden in the liver through the production of pathogenic levels of IFNγ that impede macrophage activity [13]. Conversely, the role of staphylococcal SAgs during mucosal colonization is less well understood but these toxins may function as immunological checkpoints in the nares, a major site of *S. aureus* colonization [14].

In this study, we characterized three consecutive clonal complex 5 (CC5) isolates of *S. aureus* acquired from the same patient after recurrence of an orthopedic SSI. Strains were isolated at days 19, 107 and 128 from the original surgical procedure, and the two latter strains differed from the primary isolate by the presence of a SAg-encoding plasmid. Following extensive genetic and phenotypic characterization of these isolates, including experimental bacteremia experiments in mice that are sensitive to human-tropic SAg activity, our findings suggest that the SAgs contribute to persistence of *S. aureus* bacteremia associated with SSIs.

## METHODS

### Human ethics statement

*S. aureus* bacteremia patients admitted to the London Health Sciences Centre were identified following a positive bacteremia blood culture test. The *S. aureus* isolates characterized in study were obtained from Victoria Hospital, London, Ontario, Canada, and delivered to the research laboratory on clinical swabs (Starplex Scientific). Each isolate was assigned a London Health Sciences Centre-*S. aureus* Bacteremia (SAB) unique number to maintain the patient anonymity. LHSC-SAB numbers were input into an encrypted database to be used to cross-reference patient outcomes at the conclusion of the study. The study was reviewed and approved by the University of Western Ontario Health Sciences Research Ethics Board (HSREB #105167). For experiments with human peripheral blood mononuclear cell (PBMC) assays, healthy volunteers were recruited from within the Department of Microbiology and Immunology at the University of Western Ontario, and following an outline of the risks, written informed consent was given by each volunteer before each sample was taken. After sampling, blood was anonymized and no information regarding the identity of the donor, including sex and age, was retained as per the study protocol. The study was reviewed and approved by the University of Western Ontario Health Sciences Research Ethics Board (HSREB #110859).

### Bacterial strains and growth conditions

*S. aureus* strains were routinely grown aerobically at 37°C in tryptic soy broth (TSB) or brain heart infusion broth (BHI) with shaking (250 rpm) or on tryptic soy agar (TSA) supplemented with the appropriate antibiotics as needed. For select experiment, *S. aureus* isolates were spotted onto either 5% sheep blood agar or 5% casein (skim milk) plates and grown overnight to assess hemolytic or proteolytic activity, respectively. Growth curves were performed using the Biotek Synergy H4 multimode plate reader (Agilent). *Escherichia coli* XL1-blue was used for cloning purposes and was grown aerobically at 37°C with shaking (250 rpm) in Luria-Bertani (LB) broth, or on LB agar, with the appropriate antibiotics.

### Genome sequencing

Total DNA preparations from the three *S. aureus* clinical isolates (herein referred to as SAB-0429, SAB-0485, and SAB-0495) were sequenced using paired end Illumina and long read nanopore sequencing at SeqCenter, Pittsburgh, USA. Sequence data were used to generate *de novo* assemblies using SPAdes v3.15 for both chromosome and plasmid DNA and annotated using Prokka v1.12. The genome and plasmid assemblies for the isolates have been deposited at NCBI (Biosamples SAMN42466856, SAMN42466857, SAMN42466858, PQ014898 and PQ014899). A core SNP alignment was built using snippy and snippy-core v4.1 (https://github.com/tseemann/snippy) using these isolates and a small selection of other publicly available CC5 sequences from Canada, and a phylogenetic tree was constructed using FastTree v2.1.10. The presence of previously described SAgs among the genome sequence data set was established by nucleotide BLAST (blastn) as implemented in blastable (https://github.com/bawee/blastable) using a threshold of 90% of identical positions to consider a gene present.

### T cell activation assays

*S. aureus* strains were grown in TSB overnight and sub-cultured at 1% into fresh TSB for 18 hours, cells were pelleted, and supernatants were filter sterilized. PBMCs were isolated using Ficoll-HyPaque Plus gradients (GE Healthcare), washed three times, and resuspended in complete RPMI (cRPMI) containing RPMI (Invitrogen Life Technologies) supplemented with 10% fetal bovine serum (FBS) (Wisent), 2mM L-glutamine (Gibco), 1mM sodium pyruvate (Gibco), 100 μM nonessential amino acids (Gibco), 25mM Hepes (pH 7.2) (Gibco), 100 μg/mL streptomycin, 100 U/mL penicillin (Gibco) and 2 μg/mL polymyxin B (Gibco). IL-2 concentrations were determined after 18-hours by ELISA (Invitrogen).

### Proteomic analysis

Bacteria were grown in TSB overnight and sub-cultured in fresh TSB for 18 hours, cells were pelleted, and supernatants were harvested and normalized to a OD_600_ = 1.0. Extracellular proteins were precipitated using 6% trichloroacetic acid for 30 minutes on ice. Precipitated proteins were washed in acetone and resuspended in 8M urea. Resulting samples were separated on 12% acrylamide SDS-PAGE gel and analyzed by mass spectrometry. Mass spectrometry analysis was conducted at the London Regional Proteomics Centre (University of Western Ontario, London, Canada). Briefly, protein bands within a range of 15-70 kDa were excised using an Ettan Spot Picker (GE Healthcare Life Sciences) and digestion was made in-gel using a Waters MASSPrep Automated Digestor (PerkinElmer Inc). Processed samples were pre-mixed with a matrix-assisted laser desorption ionization (MALDI) matrix and spotted on MALDI target. Sample spots were analyzed using a 5800 MALDI TOF/TOF System (AB Sciex) in reflectron-positive mode, and the peptide fingerprint masses were searched against the NCBI database for Gram-positive bacteria using the MASCOT search engine. Raw mass spectra of the samples not resulting in highly confident identification were processed and compared with the raw data of the samples identified with high confidence using the Data Explorer (AB Sciex).

### Mice

Human leukocyte antigen (HLA)-DR4-IE (DRB1*0401) transgenic mice lacking endogenous mouse MHC-II on a C57BL/6 background (herein referred to as DR4-B6 mice) [15], or conventional C57BL/6 mice (herein referred to as B6 mice), were used for *in vivo* infection experiments. Mice were sex-matched for experiments and were between 8- to 12 weeks old. DR4-B6 mice were bred within a barrier facility at the University of Western Ontario and B6 were purchased from Charles River Laboratories. Animals were housed in single-sex cages to a maximum of 4 animals per cage. Mice were provided water and food *ad libitum* and appropriate environmental enrichment was provided. The animal experiments followed the Canadian Council on Animal Care Guide to the Care and Use of Experimental Animals and the protocol was approved by the Animal Care Committee at the University of Western Ontario (Animal Use Protocol #2020-061).

### Bacteremia infection model

Single bacterial colonies were grown in TSB overnight and subcultured at 1% into fresh TSB and grown to post exponential phase (∼3 to 4 hours). The bacterial pellets were washed once and resuspended in Hank’s balanced salt solution (HBSS–-Gibco) to an OD_600_ of 0.15 which corresponds to approximately 5 × 10^7^ CFU/mL. Mice were injected via the tail vein with ∼5 × 10^6^ CFU of *S. aureus* in a total volume of 100 μL. Mice were weighed and monitored daily. At 3 days post infection, mice were euthanized, and the kidneys and livers were aseptically harvested. Organs were homogenized, plated on mannitol salt agar (MSA), and incubated at 37 °C overnight. *S. aureus* colonies were enumerated the following day with a limit of detection determined to be 3 CFU per 10 μL.

Interferon-γ (IFNγ) depletion experiments were performed as previously described [13]. Briefly, mice were treated with a 250-μg dose of anti–IFN-γ (cloneXMG1.2, BioXCell), or a rat IgG1 isotype antibody control (clone HRPN), 18 hours prior to *S. aureus* infection. All antibody doses were prepared in 100 μL PBS and administered by intraperitoneal injection.

### Curing of the pIB485-like plasmid from *S. aureus* SAB-0485

To help understand the direct contribution of the pIB485-like plasmid to *S. aureus* persistence, we attempted to cure the plasmid by repeatedly growing the strains at elevated temperatures but these experiments did not result in plasmid loss. We therefore took a genetic approach to remove the plasmid where flanking regions of staphylococcal enterotoxin R (*ser*) gene were PCR amplified, ligated and cloned in pKOR1 integration plasmid within the *attP* sites of the vector as described [16]. The pKOR1::*ser* plasmid was sequenced to confirm its integrity (Plasmidsaurus) and was subsequently electroporated into competent SAB-0485 as described [17]. Following integration of pKOR1::*ser* into the pIB485-like plasmid, to select against strains harboring the pIB485-like plasmid containing the integrated pKOR::*ser* plasmid, bacteria were treated for 3 days with anhydrotetracycline (1μg/ml) to induce the anti-sense *secY* counter selection encoded within pKOR1 [16], with subculturing of bacteria in fresh medium with the supplement every day. The final culture was serially diluted and incubated on TSA plates overnight. The colonies were first screened for absence of chloramphenicol resistance from pKOR1 and for absence of ampicillin resistance from pIB485-like. The plasmid-cured antibiotic sensitive strain was confirmed by PCR.

### Statistical analyses

Statistical analyses were performed using GraphPad Prism 10. A *p* value equal or lower than 0.05 was considered to be statistically significant. The bacterial burden calculated in the animal experiments were analyzed using the Kruskal-Wallis test with an uncorrected Dunn’s test for multiple comparisons or Mann-Whitney test.

## RESULTS

### Patient history

A patient in their 70s was admitted to London Health Science Center and received a total right knee arthroplasty, patellaplasty, and bone grafting of the distal femur on the right side to treat osteoarthritis. Following a successful surgery, the site became infected 19-days post treatment with a *S. aureus* positive blood culture (strain SAB-0429), and a second surgery was performed to remove infected tissue. The patient received cefazolin and vancomycin after the second surgery. The patient re-presented at the clinic 107-days post initial surgery with an infection in the right knee, again positive for *S. aureus* (strain SAB-0485). The patient was treated with vancomycin and sulfamethoxazole and trimethoprim (Septra). An additional sample at the right knee was collected 118 days after the initial surgery that was positive for *S. aureus* (strain SAB-0495). The patient’s antibiotic regimen was changed to cefazolin, cloxacillin and vancomycin. After this round of treatment, no further cultures of *S. aureus* were recorded.

### Strain analysis indicates the patient was infected with 2 distinct clones of *S. aureus*

We next subjected the three SAB isolates to whole genome sequencing and created *de novo* assemblies of each of the isolates to determine genetic relationships. *In silico* multi-locus sequence type (MLST) analysis indicated that all three isolates were sequence type (ST) 5 (**Table S1**) belonging to clonal complex (CC) 5. Using a selection of publicly available CC5 sequences from Canada, we determined that despite being of the same clonal lineage, these isolates were not closely related, and the SAB-0429 isolate was in a distinct clade from the SAB-0485 and SAB-0495 isolates (**Figure 1*A***). Further using the Comprehensive Antibiotic Resistance Database (CARD) [18], we were able to confirm that SAB-0429 was an MRSA encoding the Staphylococcal Cassette Chromosome *mec* (SCC*mec*) element (**Table S2**). Curiously, the CARD analysis also identified that the SAB-0485 and SAB-0495 isolates encode the BlaZ beta lactamase, but this was not present in the SAB-0429 isolate (**Table S2**). These data indicate that the patient was infected first with a clone of CC5 MRSA that was supplanted by a different CC5 MSSA.

**Figure 1.**
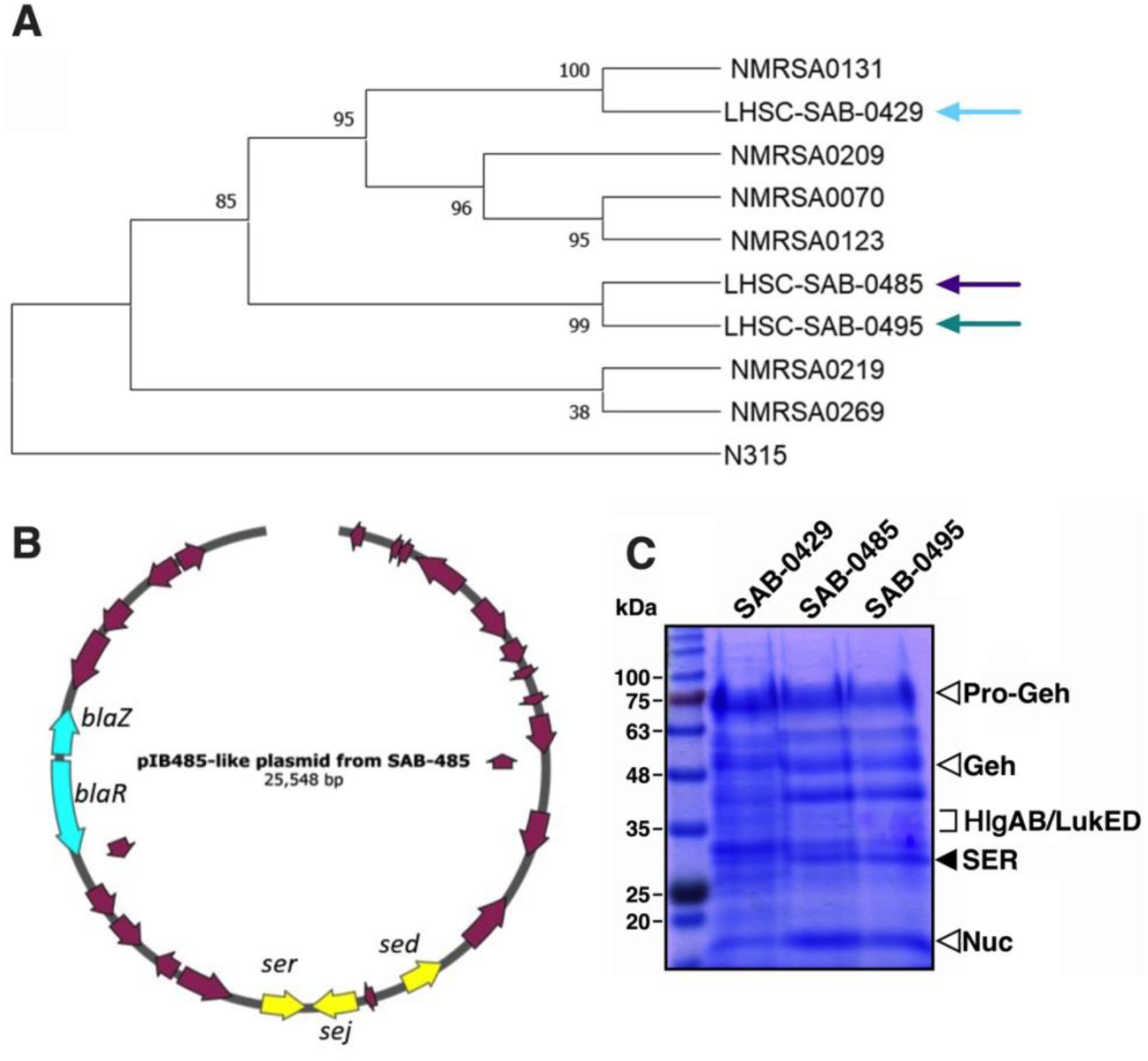
The initial methicillin-resistant *S. aureus* SAB-0429 isolate is genetically distinct from methicillin-sensitive *S. aureus* SAB-0485 and SAB-0495 isolates. (*A*) Phylogenetic tree of *S. aureus* CC5 strains including isolates SAB-0429, SAB-0485 and SAB-0495. Core SNPs (single nucleotide polymorphisms) were identified using Snippy in the three patient isolates and a small selection of other publicly available CC5 sequences from Canada. A maximum likelihood phylogeny was constructed from the aligned SNPs using FastTree v2,1,10. (B) The sequence of the pIB485-like plasmid found in both *S. aureus* SAB-0485 and SAB-0495 is presented with its main characteristics (e.g. *bla* genes and superantigen-encoding genes). (*C*) Exoprotein profiles from S. aureus SAB-0429, SAB-0485 and SAB-0495 visualized on SDS-PAGE. Open arrows indicated virulence factors found in all three isolates and the solid arrow indicates SER that was only found in SAB-0485 and SAB-0495.

### SAB-0485 and SAB-0495 have a unique set of SAgs encoded on pIB485-like plasmid

To evaluate if these strains had any difference in pathogenic potential, we assessed the genome sequencing data to determine the virulence factor composition present in these isolates. In specific reference to SAg genes, all three isolates were found to encode the staphylococcal enterotoxin genes *seg*, *sei*, *selM*, *selN*, *selW* and *selX*; however the *sed*, *sej* and *ser* genes were only in isolates SAB-0485 and SAB-0495. These three SAgs have usually been found encoded together within pIB485-like plasmids [19,20] and the presence of this plasmid in the two MSSA isolates was confirmed by assembling the plasmid from the combined long and short sequencing using *de novo* assembly (**Figure 1*B***).

To evaluate if one or all three SAgs were being expressed by SAB-0485 and SAB-0495, the extracellular proteins produced in bacterial culture from the three bacterial strains were subjected to proteomic analysis. First, SDS-PAGE analysis confirmed that the secreted profiles between the MSSA and MRSA isolates were quite different. This was further confirmed by mass spectrometry analysis (**Table S3**). SAB-0429 produced several toxins including the pro and mature forms of glycerol-ester hydrolase (Geh), nuclease (Nuc), leukocidins (LukED) and gamma hemolysins (HlgAB) (**Figure 1*C***); however, no SAg peptides were detected from *S. aureus* SAB-0429. However, from SAB-0485 and SAB-0495 supernatants, the SAg SER was identified (solid arrow, **Figure 1*C***). Notably, we also detected α-hemolysin (Hla) by mass spectrometry from both SAB-0485 and SAB-0495, but not SAB-0429 (**Table S3**).

### SAB-0485 and SAB-0495 produce increased levels of superantigen activity

To better understand how *S. aureus* may have persisted during infection, the three isolates were subjected to a range of phenotypic tests. First, the growth profile of all three isolates were assessed in rich bacterial media (tryptone soy broth) and the strains grew similar (**Figure 2*A***). To evaluate if differences in cytolytic toxin expression had an impact, hemolytic profiles of three isolates were determined by spotted the strains on TSA agar containing 5% sheep blood. Hemolysis was observed for all three isolates although this activity was decreased from the SAB-0429 isolate compared to the SAB-0485 and SAB-0495 isolates (**Figure 2*B***), consistent with an apparent reduced capacity to produce Hla (**Table S3**). In parallel, the isolates were spotted on 5% casein hydrolysis plates (skim milk plates) to examine protease activity although no differences in proteolytic activity were observed (**Figure 2*B***). Next, to evaluate if the presence of the additional plasmid encoded SAgs correlated with an increased ability to induce higher levels of T cell activation, we incubated primary human PBMCs with filter sterilized supernatant from each isolate and measured the production of IL-2. Compared to SAB-0429, both SAB-0485 and SAB-0495 were able to consistently induce T cell activation at lower supernatant dilutions, indicating that the presence of the pIB485-like plasmid correlated with higher superantigenic capacity in the two MSSA isolates (**Figure 2*C***). Taken together, the genetic and phenotypic analyses suggest that antibiotic treatment for the initial MRSA infection with SAB-0429 was successful, although the patient became reinfected with a persistent MSSA isolate that produced larger amounts of virulence promoting toxins.

**Figure 2.**
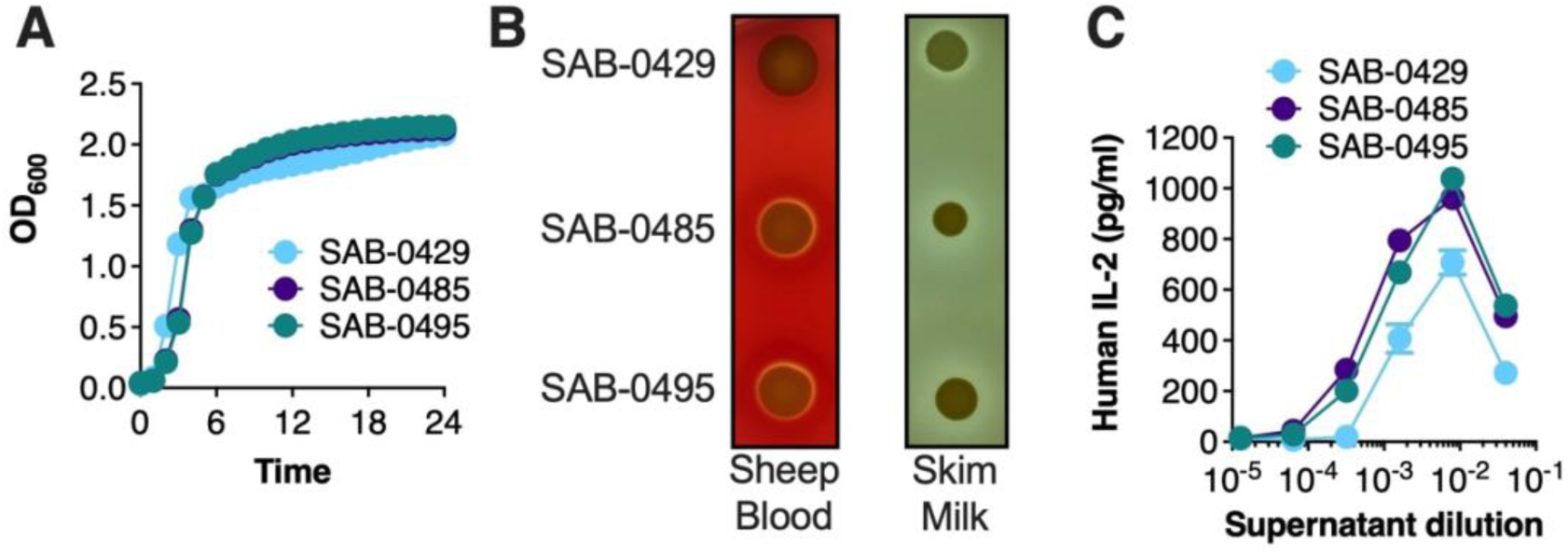
Phenotypic analyses demonstrate enhance superantigen activity from S. aureus SAB-0485 and SAB-0495 compared to SAB-0429. (*A*) The three *S. aureus* patient isolates were grown in TSB for 24 hours with agitations in a multimode plate reader and their optical density at 600 nm was monitored every hour. Each data point represents the mean of three independent experiments. (*B*). Hemolytic and proteolytic activity of each isolate assessed on 5% sheep blood TSA plates and skim milk agar plate, respectively. The image is a representative image of experiments replicated at least three times. (*C*) IL-2 production from supernatants from each isolate grown in TSB for 18 hours. Supernatants were filter-sterilized before exposure to human PBMCs and IL-2 concentrations were measured by ELISA. Each data point represents the mean ± SEM of three independent experiments using different donors.

### Isolates containing the pIB485-like plasmid persist at higher level in the liver

The increased superantigen activity of the MSSA isolates (**Figure 2*C***) may have promoted persistence of these strains following the second surgery and the relapse of infection in the patient’s knee. To evaluate if the SAB-0485 and SAB-0495 strains exhibited increased persistence *in vivo* compared to the SAB-0429 isolate, we utilized an experimental model of bacteremia in transgenic mice that express human MHC-II molecules (DR4-B6) and are sensitive to SAg function [13,21]. The susceptibility of the DR4-B6 strain to exoproteins secreted by each *S. aureus* strain was tested by exposing extracted splenocytes with isolate supernatants and assessing for IL-2 production. Supernatants from both SAB-0485 and SAB-0495 resulted in an increased production of IL-2 compared to SAB-0429, especially visible at the dilution factor of ∼1/250, noting that the more concentrated supernatants contain active cytolytic toxins that kill the immune cells (**Figure 3*A***). To identify pathogenic differences between these strains during experimental bacteremia, DR4-B6 mice were intravenously inoculated with the different *S. aureus* isolates and bacterial burden in the liver and kidneys were assessed 3 days post-inoculation. The bacterial burden in the liver of DR4-B6 mice was increased for both SAB-0485 and SAB-0495 compared with SAB-0429 (**Figure 3*B***), although there were no differences in bacterial counts in the kidneys (**Figure 3*C***). These data suggest that the SAgs encoded on the pIB485-like plasmid may be involved in promoting the persistence of the latter SAB-0485 and SAB-0495 isolates compared with the initial SAB-0429 isolate during bloodstream infection.

**Figure 3.**
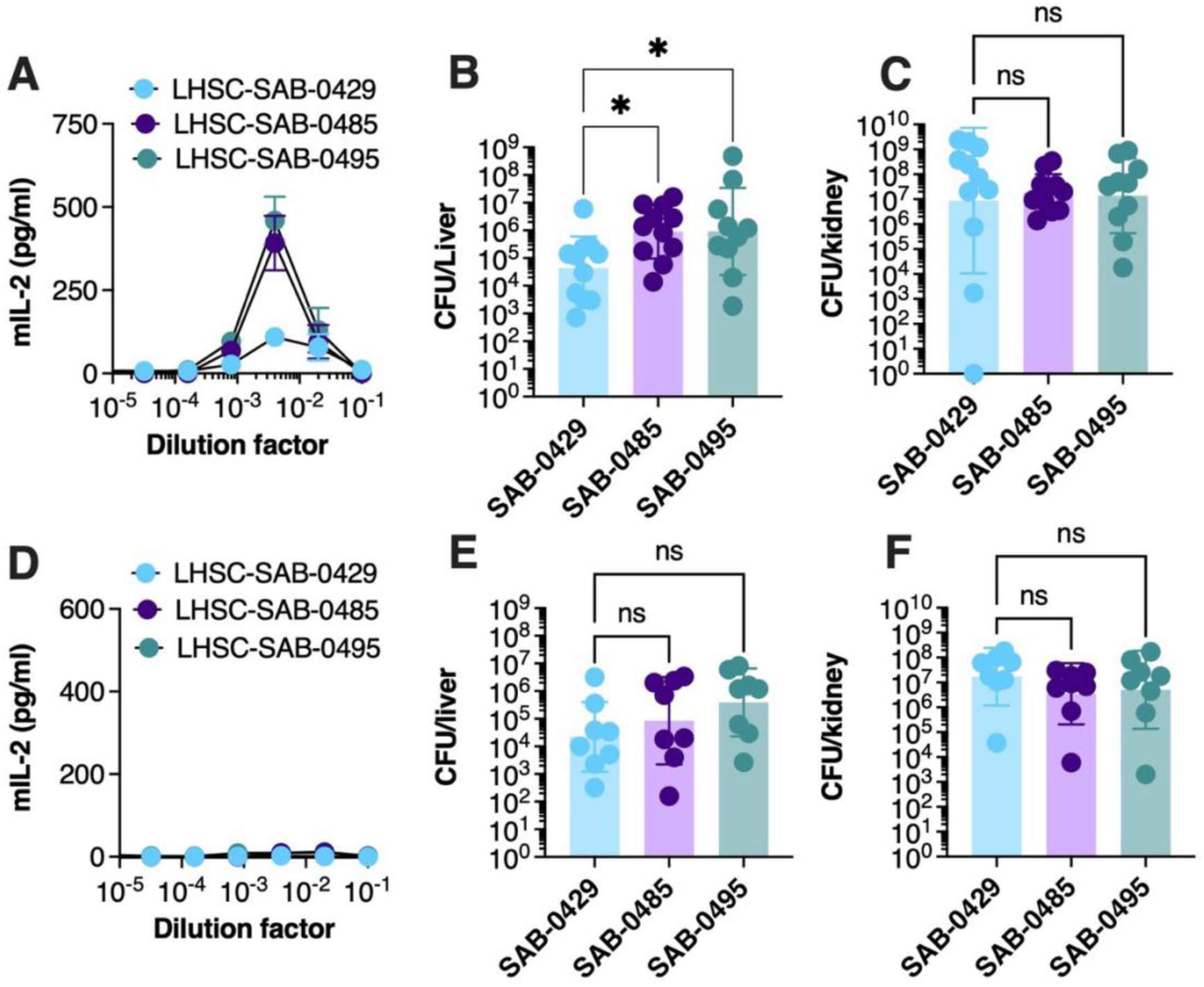
*S. aureus* isolates SAB-0485 and SAB-0495 containing the pAB485-like plasmid persist at a higher bacterial counts compared to SAB-0429 in a SAg-sensitive model of bacteremia. DR4-B6 (*A*) or B6 (*D*) splenocytes were isolated and challenged against supernatant issued from each bacterial isolate and IL-2 production from stimulated cells was measure by ELISA. Data presented is the mean of three independent experiments ±SD. Each isolate was intravenously injected at 5 × 10^7^ CFU/mL to DR4-B6 (B&C) or B6 (E&F) animals and bacterial burden of both kidneys (B&E) and liver (C&F) was assessed 3 days post-infection. Each dot represents one animal. The results are represented as the geometric mean of at least 8 biological replicates ± geometric SD. Significant differences were determined using the Kruskal–Wallis test with the uncorrected Dunn’s test for multiple comparisons (*P < 0.05).

Hla has also been implicated in liver persistence by *S. aureus* [15]. As this toxin showed increased expression from the proteomic analysis (**Table S3**) and that phenotypic assessment showed decreased hemolytic activity for SAB-0429 (**Figure 2*B***), it was important to evaluate a potential role of Hla in the bacteremia model. Conventional mouse models are sensitive to the activity of this toxin [22]; therefore, we repeated our bacteremia analysis in conventional B6 mice. Splenocytes from conventional B6 mice were first co-incubated with supernatants from each isolate which did not induce any detectable T cell activation, confirming the lack of susceptibility of these mice to SAg (**Figure 3*D***). Next, the experimental bacteremia experiment was repeated in conventional B6 mice and although there was a trend suggesting increased bacterial recovery from the liver for the two MSSA isolates, this was not statistically different (**Figure 3*E***). As with the DR4-B6 mouse experiments, there were no obvious differences in bacterial counts recovered from the kidneys. These collective data suggest that the differences we observed between the tested *S. aureus* strains was due primarily to the additional SAgs encoded by SAB-0485 and SAB-0495, rather than the expression of other virulence factors.

### Loss of pIB485-like decreases the bacterial burden in the liver

To determine if the increased bacterial burden within the liver of DR4-B6 mice was due to the set of SAgs encoded within pIB485-like plasmid, the cured this plasmid from SAB-0485 and this strain was evaluated using the bacteremia model in DR4-B6 mice. First, the supernatants from either wildtype *S. aureus* SAB-0485 or the isogenic strain lacking the pIB485-like plasmid were tested for IL-2 production from DR4-B6 splenocytes and a decreased stimulation of PBMC was observed with the plasmid cured strain (**Figure 4*A***). We hypothesized that this decrease in T cell stimulation would correlate with decreased bacterial burden within the liver in the DR4-B6 bloodstream infection model. Indeed, in absence of the pIB485-like plasmid, this strain reached a lower bacterial burden in the liver of the mice compared to wild-type SAB-0429 (**Figure 4*B***).

**Figure 4.**
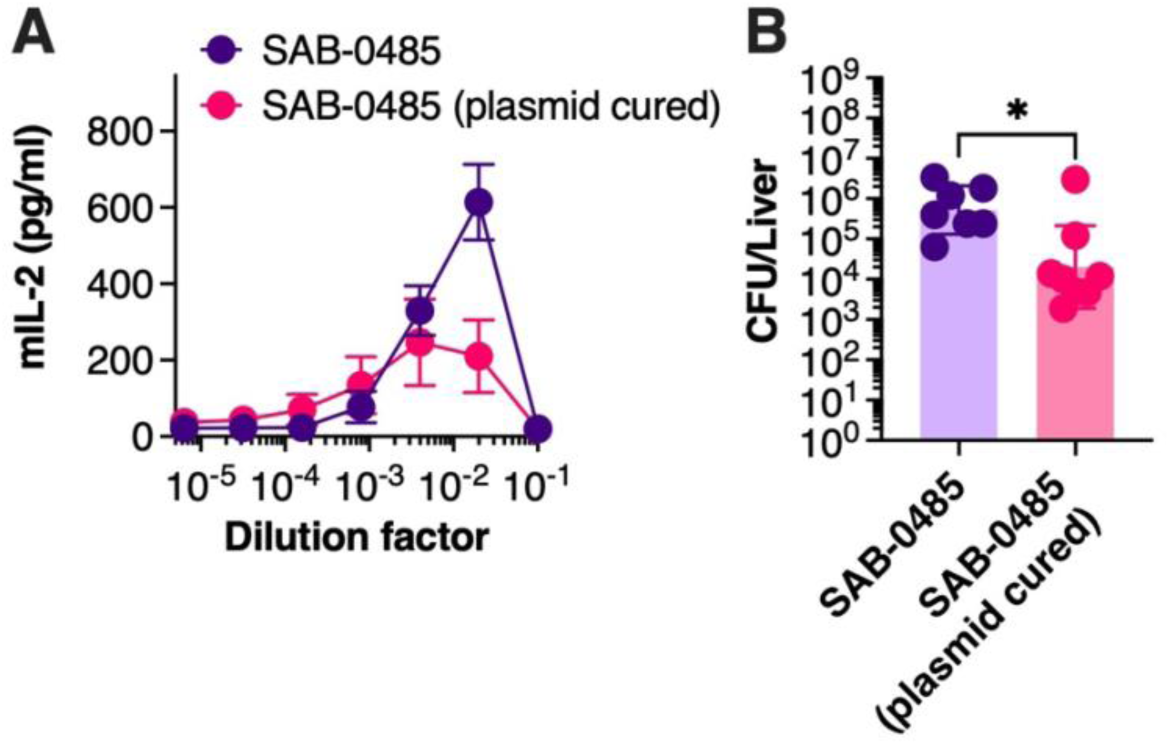
Loss of the pIB485-like plasmid in the SAB-0485 isolate demonstrates a decreased bacterial burden in the liver. (*A*) B6-DR4 splenocytes were isolated and challenged against supernatant issued from each bacterial strain (SAB-0485 or SAB-0485 without plasmid). The graph represents murine IL-2 quantification during at least three independent experiments. (*B*) The same strains were intravenously injected to DR4-B6 animals to perform the bacteremia model and the bacterial burden in the liver is represented as the geometric mean of at least 7 animals ± geometric SD. Each dot represents one mouse. Significant differences were determined using Mann Whitney test (*P < 0.05).

### pIB485-like encoded SAgs promote a pathogenic IFNγ response

We previously demonstrated that SAgs could promote a pathogenic IFNγ response that modifies macrophage responses and allows *S. aureus* to persist and replicate more effectively within macrophages [13]. This mechanism could potentially explain why the two MSSA isolates persisted for longer in the patient compared with the MRSA clone despite antibiotic treatment. To test this hypothesis, we utilized an IFNγ depletion protocol in DR4-B6 mice prior to infection with either SAB-0429 or SAB-0485, and bacterial liver and kidney burdens were enumerated at 72 hours post-infection (**Figure 5*A***). For infections with *S. aureus* SAB-0429, depletion of IFNγ had no measurable impact on bacterial recovery from the liver or kidneys compared to the isotype antibody treatment control. For animals infected with *S.* aureus SAB-0485, IFNγ depletion resulted in a significant reduction in bacterial recovery from the liver (**Figure 5*B***) but again did not alter kidney burden (**Figure 5*C***). Importantly, the IFNγ depletion for the SAB-0485 isolate reduced bacterial recovery in the liver that was equivalent to the SAB-0429 isolate, indicating that removing the pathogenic IFNγ production mitigated the activity of the plasmid encoded SAgs. Together this demonstrates that the pIB485-like encoded SAgs can promote excessive IFNγ production and promote bacterial persistence of these isolates during experimental bacteremia.

**Figure 5.**
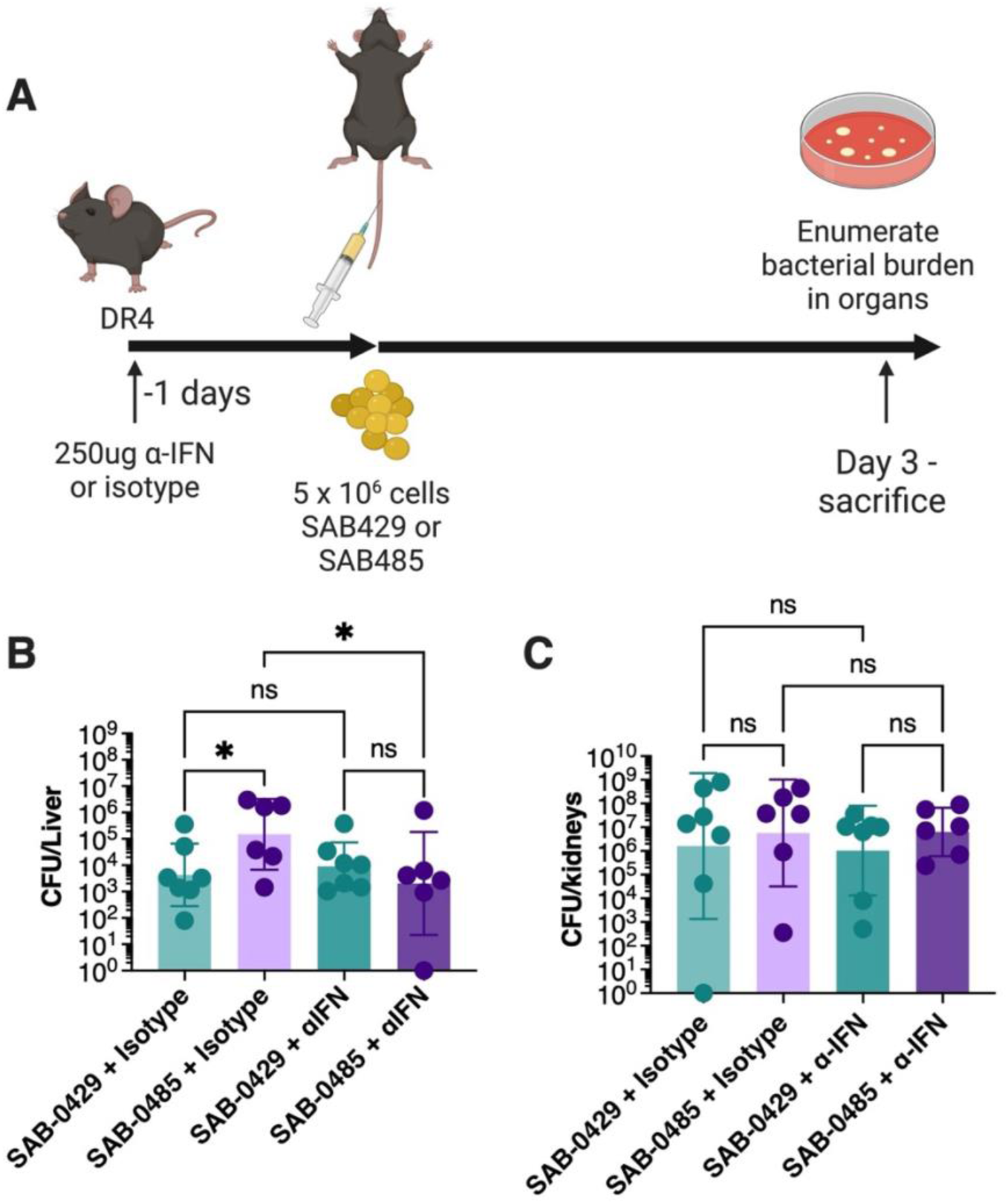
Depletion of Interferon gamma eliminates SER-mediated bacterial persistence during *S. aureus* bacteremia. (*A*) The bacteremia model was repeated with the isolates SAB-0429 and SAB-0485 after intraperitoneally injecting the animal with either depleting antibodies for IFNγ or an isotype control 18 hours before *S. aureus* intravenous challenge. Both the kidneys (*B*) and the liver (*C*) were collected and homogenized for bacterial burden. The results are represented as the geometric mean of at least 8 biological replicates ± geometric SD. Significant differences were determined using the Kruskal– Wallis test with the uncorrected Dunn’s test for multiple comparisons (*P < 0.05).

## Discussion

*S. aureus* infections acquired in the hospital setting can result in life-threatening disease and poor outcomes which may be further complicated by antimicrobial resistance. In addition, microbial mechanisms that promote persistence during infection can further exacerbate disease. In this work, we encountered MSSA isolates of *S. aureus* that were able to persist in a patient despite antimicrobial therapy. Based on comparisons with the earlier MRSA isolate from the same patient, and combined with experimental infection models, we were able to determine that the increased persistence of these MSSA strains was associated with the production of plasmid-encoded SAg toxins. We initially considered that this may be an example of in-patient evolution, but genome sequencing determined that although the main differentiating determinant of the strains was the pIB485-like plasmid, the SAB-0429 isolate was not ancestral to the SAB-0485 and SAB-0495 isolates (**Figure 1*A***). Therefore, this patient may have been colonized simultaneously at the injury site with different *S. aureus* clones or was subsequently infected during the period between presentations at the clinic.

All three *S. aureus* isolates were identified to be members of the CC5 clade. This clonal complex is a clinically important group and frequently isolated from nosocomial infections. In Ontario and Canada, CC5 has been related with community-associated infections, especially in the case of MRSA isolates [23]. Strains from CC5 also harbor high variability in their SAg complement, and this can include the presence or absence of the pIB485 -like plasmid in both MRSA and MSSA lineages of this clonal complex [23]. Indeed, the pIB485-like plasmid contributed to this genetic variability between the MRSA isolates in this study. Interestingly, multiple experiments growing S. aureus SAB-0485 at elevated temperatures did not result in curing of this plasmid, suggesting this is a stable genetic element in *S. aureus* that may be associated with persistent infection and worse patient outcome.

The MSSA isolates we identified were able to survive in the patient in between disease episodes and cause resurgence of bacteremia despite high levels of antibiotic stress. Subsequently, we were able to demonstrate that the MSSA isolates were persisting in experimental *in vivo* bacteremia model, in part, by inducing a pathogenic IFNγ response (**Figure 5**). We previously demonstrated that this mechanism could support the replication of *S. aureus* inside macrophages and allow the bacteria to persist *in vivo*. Further, being able to persist and replicate within a macrophage may effectively contribute to avoidance of killing by antibiotics [13]. Other mechanisms of persistence within macrophages have been previously described for many bacterial pathogens; however these mechanisms usually include a ‘persister-type’ phenotype [24,25]. In the current study, *S. aureus* persistence may allow survival of the bacterium independent of this persister phenotype as the quick recovery of bacteria following the organs collection suggests that *S. aureus* was in a replicative state in the animal. Although both SAB-0485 and SAB-0495 harbored several SAg genes, only SER was detected from *in vitro* conditions by proteomics, and we demonstrated that this impacted the bacterial burden within the liver during bacteremia. Altogether, SER demonstrated similar characteristics to SEA [21], SEB and SEC [13], and SE*l*-W [26] produced from other *S. aureus* strains in bloodstream infections by enhancing persistence in the liver.

Due to limitations in sampling, we were unable to fully establish how this patient experienced infections with two distinct clones of *S. aureus*. Samples at admission were not retained so we were unable to assess if this patient was nasally colonized prior to infection, and if this was the source of the surgical complication. During the disease, both co-infection with the MSSA and MRSA clones is possible as well as hematological spread from another source. In both scenarios, a virulence mechanism that enhanced survival in macrophages would have provided a selective advantage for the MSSA strains in the patient and may have led to the clearance of the MRSA clone.

This study highlights how virulence factors may provide an evolutionary advantage to *S. aureus* strains in certain patient settings. Beyond classical antimicrobial resistance mechanisms, *S. aureus* is also potent at manipulating its environment and shielding itself from the immune system. Although these findings do not diminish the concerns around antimicrobial resistance, this research demonstrates the importance of considering the niche of *S. aureus,* the range of virulence factors encoded in its genome, and factors that are produced by the bacterium during infection that contribute to disease.

## Data Availability

All data produced in the present work are contained in the manuscript.

