## Supplementary material for "Staphylococcal superantigens promote bacterial persistence following postoperative surgical site infection": Tables S1-S3

**Table S1. MLST and Spa typing for the *S. aureus* strains isolated from the same bacteremia patient.**

| | Allelic Profile | | | | | | | MLST | Spa |
|---|---|---|---|---|---|---|---|---|---|
| | *arcC* | *aroE* | *glpF* | *gmK* | *pta* | *tpi* | *yquiL* | | |
| SAB-0429 | 1 | 4 | 1 | 4 | 12 | 1 | 10 | ST5 | t002 |
| SAB-0485 | 1 | 4 | 1 | 4 | 12 | 1 | 10 | ST5 | t002 |
| SAB-0495 | 1 | 4 | 1 | 4 | 12 | 1 | 10 | ST5 | t002 |

**Table S2. Antibiotic resistance genes from genomic analyses for the *S. aureus* strains isolated from the same bacteremia patient..**

| | | % identity | | |
|---|---|---|---|---|
| Gene | AMR Gene Family | SAB-0429 | SAB-0485 | SAB-0495 |
| *mecR1* | Methicillin resistant PBP2 | 100 | 0 | 0 |
| *mecI* | Methicillin resistant PBP2 | 100 | 0 | 0 |
| *mecA* | Methicillin resistant PBP2 | 99.7 | 0 | 0 |
| blaZ | β-lactamase | 0 | 93.95 | 93.95 |

**Table S3. Proteomic analysis of the supernatants from the *S. aureus* strains isolated from the same bacteremia patient.**

| Spot # | Predicted Size | Protein hit | P-value |
|---|---|---|---|
| 0429-15 | 23949 | Thermonuclease [*S. aureus* subsp. *aureus* CIG1750] | 6,90E-08 |
| 0429-20 | 18358 | Alkyl hydroperoxide reductase, partial [*S. aureus*] | 2,20E-05 |
| 0429-25 | 22375 | Superantigen-like protein (Sav0433) from *S. aureus* Mu50 | 6,90E-07 |
| 0429-28 | 32220 | Glycerophosphodiester phosphodiesterase, partial [*S. aureus*] | 2,20E-08 |
| | 34968 | Gamma-hemolysin subunit A [*S. aureus*] | 1,40E-07 |
| 0429-35 | 34028 | Chain A, leukocidin F (Hlgb) from *S. aureus* | 5,50E-05 |
| | 32090 | Pyridoxal biosynthesis protein [*S. aureus*] | 0,0035 |
| | 31410 | Leukotoxin LukD [*S. aureus* DAR5844] | 1,10E-08 |
| | 27094 | Gamma-hemolysin protein B [*S. aureus*] | 2,80E-07 |
| 0429-70 | 71387 | Lipase [*S. aureus*] | 1,70E-11 |
| 0485-15 | 25216 | Thermonuclease [*S. aureus*] | 4,40E-08 |
| 0485-20 | 10311 | 50S ribosomal protein L6, partial [*S. aureus* M0271] | 0,00087 |
| 0485-25 | 30023 | Enterotoxin [*S. aureus*] (SER) | 1,40E-05 |
| 0485-28 | 32220 | Glycerophosphodiester phosphodiesterase [*S. aureus*] | 6,90E-07 |
| | 33115 | Chain B of octameric pore of gamma-hemolysin from *S. aureus* | 5,50E-05 |
| | 36389 | Leukotoxin LukE [*S. aureus* LPIH6021] | 8,70E-06 |
| | 36349 | Gamma-hemolysin component A [*S. aureus* VRS2] | 0,00014 |
| 0485-30 | 34437 | Chain A, crystal structure of alpha-hemolysin | 4,40E-11 |
| 0485-35 | 34028 | Chain A, leukocidin F (Hlgb) from *S. aureus* | 1,40E-09 |
| | 36429 | Gamma-hemolysin component B [*S. aureus* CIG1835] | 3,50E-09 |
| | 31410 | Leukotoxin LukD [*S. aureus* DAR5844] | 2,20E-08 |
| | 35244 | Chain A, of octameric pore of gamma-hemolysin from *S. aureus* | 3,50E-06 |
| 0485-70 | 76495 | Lipase [*S. aureus*] | 8,70E-21 |
| 0495-15 | 23949 | *S. aureus* sub. sp. *aureus* thermonuclease | |
| 0495-20 | 12679 | *S. aureus* 50s ribosomal protein L6 | |
| 0495-25 | 30023 | Enterotoxin (SER) | 0,00011 |
| 0495-28 | 32220 | Glycerophosphodiester phosphodiesterase [*S. aureus*] | 1,40E-10 |
| | 33115 | Chain B of octameric pore form of gamma-hemolysin, *S. aureus* | 2,20E-06 |
| 0495-30 | 34437 | Chain A of Alpha-Hemolysin | 2,80E-08 |
| 0495-35 | 34028 | Chain A, leukocidin F (Hlgb) from *S. aureus* | 1,70E-05 |
| | 36698 | Gamma-hemolysin subunit B [S. aureus] | 5,50E-05 |
| | 34870 | Chain B, covalent S-f heterodimer of Gamma-hemolysin | 0,00069 |
| | 31410 | Leukotoxin LukD [*S. aureus* DAR5844] | 2,00E-03 |
| | 35244 | Chain A, of octameric pore of gamma-hemolysin From *S. aureus* | 9,10E-03 |
| | 27094 | Gamma-hemolysin protein B [*S. aureus*] | 1,30E-02 |
| 0495-70 | 76486 | Lipase [*S. aureus*] | 5,50E-14 |
